# Identification of *Schistosoma haematobium* and *Schistosoma mansoni* linear B-cell epitopes with diagnostic potential using *in silico* immunoinformatic tools and peptide microarray technology

**DOI:** 10.1101/2023.12.28.23300599

**Authors:** Arthur Vengesai, Marble Manuwa, Herald Midzi, Masimba Mandeya, Victor Muleya, Keith Mujeni, Isaac Chipako, Dean Goldring, Takafira Mduluza

**Author notes:** Corresponding author, (AV).

## Abstract

**Introduction:** Immunoinformatic tools can be used to predict schistosome-specific B-cell epitopes with little sequence identity to human proteins and antigens other than the target. This study reports an approach for identifying schistosome peptides mimicking linear B-cell epitopes using in-silico tools and peptide microarray immunoassays validation.

**Method:** Firstly, a comprehensive literature search was conducted to obtain published schistosome-specific peptides and recombinant proteins with the best overall diagnostic performances. For novel peptides, linear B-cell epitopes were predicted from target recombinant proteins using ABCpred, Bcepred and BepiPred 2.0 *in-silico* tools. Together with the published peptides, predicted peptides with the highest probability of being B-cell epitopes and the lowest sequence identity with proteins from human and other pathogens were selected. Antibodies against the peptides were measured in sera, using peptide microarray immunoassays. Area under the ROC curve was calculated to assess the overall diagnostic performances of the peptides.

**Results:** Peptide AA81008-19-30 had excellent and acceptable diagnostic performances for discriminating *S. mansoni* and *S. haematobium* positives from healthy controls with AUC values of 0.8043 and 0.7326 respectively for IgG. Peptides MS3_10186-123-131, MS3_10385-339-354, SmSPI-177-193, SmSPI-379-388, MS3-10186-40-49 and SmS-197-214 had acceptable diagnostic performances for discriminating *S. mansoni* positives from healthy controls with AUC values ranging from 0.7098 to 0.7763 for IgG. Peptides SmSPI-359-372, Smp126160-438-452 and MS3 10186-25-41 had acceptable diagnostic performances for discriminating *S. mansoni* positives from *S. mansoni* negatives with AUC values of 0.7124, 0.7156 and 0.7115 respectively for IgG. Peptide MS3-10186-40-49 had an acceptable diagnostic performance for discriminating *S. mansoni* positives from healthy controls with an AUC value of 0.7413 for IgM.

**Conclusion:** One peptide with a good diagnostic performance and 9 peptides with acceptable diagnostic performances were identified using the immunoinformatic approach and peptide microarray validation. There is need for evaluation with true negatives and a good reference.

**Author summary:** Schistosomiasis commonly known as bilharzia is the third most significant tropical disease after malaria and soil-transmitted helminthiases. Like other neglected tropical diseases common in Zimbabwe, schistosomiasis remains mostly undiagnosed or undetected. This is partly due to the fact that reliable identification of parasites requires expertise for specimen preparation, and microscopic examination which are largely unavailable in most rural clinics. This limitation is further compounded by the fact that the recommended microscopy-based methods for schistosomiasis diagnosis lack sensitivity, especially in infections of low intensity. To overcome some of the caveats associated with microscopy-based methods, highly sensitive serological tests have been utilized. Unfortunately, currently available serological tests have low specificity and show cross-reactivity with other helminthic infections. One way to mitigate the cross-reactivity challenge and increase the specificity, is to use immunoinformatic tools and immunoassays to identify schistosomiasis species-specific immunogenic peptides mimicking B-cell epitopes (short amino acid sequences of the antigen that reacts with antibodies). Utilizing immunoinformatic tools coupled with peptide microarray immunoassay validation approach several peptides that can be used to develop diagnostic tools for showing exposure to infection for people living in non-endemic or low-transmission areas were identified in the current study.

## 2 Introduction

Schistosomiasis is a neglected tropical disease caused by blood flukes from the genus *Schistosoma* (1). Zimbabwe is endemic to urogenital schistosomiasis caused by *Schistosoma haematobium* and intestinal schistosomiasis caused by *Schistosoma mansoni* (2–4). Besides effective implementation of mass drug administration (MDA) campaigns, access to safe water, improved sanitation and snail control, diagnostic tests are important tools for achieving and sustaining schistosomiasis elimination (1,5,6). Diagnostic tests are important for schistosomiasis surveillance and control. They play a vital role in guiding the distribution of current program resources and the implementation and evaluation of schistosomiasis intervention strategies (5,6).

The recommended method for schistosomiasis diagnosis is the detection of schistosome-specific eggs in stool or urine specimens by microscopy. For urogenital schistosomiasis, the urine filtration technique is the standard diagnostic method and for intestinal schistosomiasis, the Kato Katz technique is the standard diagnostic method (6–8). In addition to high specificity (7,9), both techniques have minimal operational costs (the test kits are inexpensive), low complexity and they are relatively easy to perform in resource-limited field settings (6,8,9). Moreover, prepared Kato Katz slides can be stored for months at room temperature for later microscopic examination.

Nevertheless, both techniques have significant disadvantages including, the need for qualified personnel to prepare and examine slides, poor reproducibility and most importantly low sensitivity (7,8,10,11). The sensitivity of the techniques is limited by the host infection intensity, daily variation of schistosome egg excretion and uneven distribution of eggs within stool specimens (11). Additionally, both methods are unable to diagnose recent infections (for instance, in cases where worms have not yet started to produce eggs) or single worm/sex infections and the Kato Katz technique is unable to analyse watery stool specimens (9). The lack of sensitivity especially in low endemic areas and after successful control interventions lead to underestimation of true schistosomiasis prevalence in such settings (7–10). Moreover, undetected and untreated individuals may continue schistosomiasis transmission by contaminating fresh water sources with urine and faeces containing schistosome eggs (1,9). Due to the numerous disadvantages associated with the microscope based techniques, the use of other methods like molecular detection, circulating cathodic antigen (CCA), circulating anodic antigen (CAA) has gathered pace (8). These alternative methods for schistosomiasis diagnosis have their own advantages and limitations too.

Molecular methods targeting DNA of the parasite are more sensitive and specific and can detect early stage schistosome infections (8). However, molecular methods require skilled laboratory personnel, expensive and fragile equipment and they are time consuming, thus impeding their use as point-of-care tools (POC) in remote resource-limited endemic areas (5). Although CCA and CAA allow for rapid schistosomiasis diagnosis, these methods have their own shortcomings (3,7,8). The CCA test is sensitive for moderate to high level *S. mansoni* infections but not for *S. haematobium* infections, and its widespread use in poor rural endemic areas may be limited by its cost, currently around US $1.75 per test (3,12,13). While the CAA test is more sensitive than the CCA, it is labour intensive and riddled with a complicated assay procedure. This major drawback is further compounded by the fact that there has not been any commercially available CAA tests to date (8).

To overcome some of the drawbacks associated with microscopy, molecular, CCA and CAA based diagnostic tests; serological tests have been utilized (14). The use of serological tests has gained traction due to higher sensitivities compared to microscopy based techniques (8). However, currently available serological tests exhibit low specificity and are laden with cross-reactivity issues with other helminthic infections due to shared antigenic epitopes (14,15). One way to mitigate the cross-reactivity challenge and increase specificity is to use bioinformatic and proteomics tools to predict schistosome specific immunodominant B-cell epitopes with little or no sequence identity to proteins other than the target (4,14,16). However, to date, only a limited number of linear B-cell epitopes have been identified for the serological diagnosis of *S. haematobium* and *S. mansoni* (17–19). It is against this background that we present an approach for predicting schistosome specific peptides mimicking linear B-cell epitopes using *in-silico* tools and peptide microarray technology. Several methods can be used for the identification and prediction of linear B-cell epitopes. As previously described, two methods *in-silico* prediction and identification of published peptides reduce the burden and costs associated with epitope mapping by decreasing the list of potential targets for experimental testing (20–24). Therefore, in the present study a comprehensive literature search was conducted to identify published schistosome-specific peptides for inclusion in the peptide microarray immunoassays.

## 3 Methods

### 3.1 Ethical approval

Approval to conduct the study was obtained from Medical Research Council of Zimbabwe (MRCZ/A/2571). Provincial Medical directors, District Medical officers, councillors, and headmen provided the permission to carry out the study in the districts. Before enrolment, the aims and procedures of the study were explained to all participants and their parents or guardians in English or Shona. Consent forms were supplied to the children’s parents or guardians for signing. Enrolment into the study was completely voluntary and parent or guardians were free to withdraw their children at any time with no need of explanation.

### 3.2 Study area and population

Children living in schistosomiasis endemic areas were purposively selected for the cross-sectional study. The children were permanent residents of Mount Darwin and Shamva rural districts located in Mashonaland central (31°40′0” E longitude and 17°10′0” S latitude) Northeast of Zimbabwe. Negative control sera were obtained from *Schistosoma* microscopy negative individuals without history of exposure or contact with *Schistosoma* contaminated water. These negative control sera were obtained from permanent residents of Glaudina a high density urban area located in Harare the capital city of Zimbabwe.

### 3.3 Blood collection

Experienced local nurses collected approximately 5 ml of venous blood for the peptide microarray serological assays from each participant. The 5ml blood limit was within the guidelines for children issued by Medical Research Council of Zimbabwe research ethics committee.

### 3.4 Parasitology examination

Urine and stool specimens were collected between 10:00 am and 14:00 pm for optimal egg passage necessary for diagnosis of schistosomiasis. The specimens were stored away from direct sunlight until processing. The urine filtration technique was used in the diagnosis of *S. haematobium.* This was repeated for three consecutive days to avoid misdiagnosis due to day-to-day egg variation. *S. mansoni* was diagnosed using the Kato Katz method and the formal ether concentration technique to improve accuracy. Participants were classified as infected if at least one parasitic egg was detected.

### 3.5 Peptide selection and *in silico* prediction

Two methods were used for the identification and prediction of linear B-cell epitopes (peptides). These were a comprehensive literature search for published synthetic peptides (25) and *in silico* prediction of novel peptides. For novel peptides, a systematic scoping review (25) was conducted to identify recombinant proteins with the best overall diagnostic performance with different serological assays including protein microarray, ELISA and POC immunochromatographic tests for *S. haematobium* and *S. mansoni*. The sequences and the alpha fold predicted structures of the identified recombinant proteins were obtained from Uniprot. SignalP 6.0 was used to identify the presence of signal peptides. Transmembrane domains were predicted using SOSUI and cellular localisation was predicted using WoLF Psort II. DeepView/Swiss-PDB Viewer (www.expasy.org/spdbv/) was used for spatial location of the candidate peptides on the recombinant protein crystal structures.

Linear B cell epitopes were predicted using three different programs namely ABCpred, Bcepred and BepiPred 2.0. ABCpred uses artificial neural network, Bcepred predicts using physico-chemical properties of amino acids and BepiPred 2.0 uses a random forest algorithm. Peptides with the lowest sequence identity with human protein and proteins from other human pathogens were then selected using the NCBI Protein BLAST to minimize cross reactivity. Peptides that had the highest probability of being B-cell epitopes and the lowest sequence identity with proteins from human and other pathogens were selected for inclusion on the peptide microarrays.

### 3.6 Peptide microarray content

The peptide microarrays were fabricated by PEPperPRINT GmbH (Heidelberg, Germany). The peptide microarrays contained 16 identical sub-arrays (copies). Each sub-array contained 122 *S. haematobium* and *S. mansoni* 9-16 amino acids long peptides printed randomly in duplicate. The peptides on each subarray were framed by HA (YPYDVPDYAG, 5 spots) and polio (YPYDVPDYAG, 3 spots) control peptides. Additionally, each sub-array was framed by glycine spacers (G spots).

### 3.7 Peptide microarrays immunoassay

Immunoassays were performed by PEPperPRINT GmbH (Heidelberg, Germany) as previously described (26,27). Briefly, the immunoassays involved two steps on the same microarray. The preincubation step which was performed to identify false positive signals by binding of the fluorescent labelled secondary antibody followed by the main incubation with serum and the secondary antibodies. At each step there was pre-swelling of the peptide microarrays with washing buffer (PBS, pH 7.4 with 0.05% Tween 20) for 10 minutes. The peptide microarrays were scanned using an LI-COR Odyssey Imaging System, scanning offset 0.65 mm, resolution 21 μm, scanning intensities of 7/7 (red = 680 nm / green = 800 nm). To ensure that all microarrays were responding correctly, all steps were repeated with the Cy3-conjugated anti-HA control antibody and Cy3-conjugated anti-polio control antibodies. Quantification of spot intensities was based on 16-bit gray scale tiff files. Microarray image analysis was done with PepSlide1 Analyzer and resulted in raw data CSV files for each sample (green = 800 nm = IgM staining, red = 680 nm = IgG staining). A PEPperPRINT software algorithm calculated averaged median foreground intensities (foreground-background signal) and spot-to-spot deviations of spot duplicates and assembled the outcome in summary files. For duplicate spots a maximum spot-to-spot deviation of 50% was tolerated, otherwise the corresponding intensity value was zeroed.

### 3.8 Identifying schistome-specific antigenic peptides

The negative cut-off was determined by averaging the negative control readings (10 sera obtained from schistosomiasis unexposed and uninfected individuals with no prior history of *Schistosoma* infections) and adding 3 standard deviations. A positive response was defined as fluorescence intensity above the negative cut-off for each specific peptide for both IgG and IgM. Peptides for which at least one infected individual was positive were selected for further analysis. Statistical comparison between groups was done by the Kruskal-Wallis equality-of-populations rank test and *p-values* less 0.05 were considered statistically significant. Diagnostic accuracy was evaluated by receiver operating characteristic (ROC) curve analysis. The area under the ROC curve (AUC) was calculated to assess the overall diagnostic performance of peptides that were able to distinguish schistosome positives from schistosome negatives or healthy controls. Data curation and analyses were performed with Microsoft excel and Stata v17 (Stata, College Station, Texas, USA) respectively.

## 4 Results

### 4.1 Demographic and parasitology characteristics

The case control diagnostic accuracy study consisted of 135 participants [62.96 % (85) females and 37.04 % (50) males] with a median age of 9 (IQR: 4-12) from Mashonaland central (92.59 %) a high schistosomiasis endemic area (2) and Harare (7.41 %). Among the participants Mashonaland central 43 were confirmed to be infected with *S. haematobium* and 36 confirmed to be uninfected with *S. haematobium* by the urine filtration technique (**S1 file**). For *S. mansoni* 46 were confirmed to be infected and 37 were confirmed to be uninfected by the Kato Katz method and the formal ether concentration technique (**S1 file**). Ten participants without confirmed *Schistosoma* infections and without a history of infection and contact with contaminated water were recruited from Glaudina a high density urban area located in Harare (**S1 file**).

### 4.2 Identification and characterisation *schistosome* recombinant proteins for novel peptides prediction

Two recombinant proteins MS3_10385 a serine protease inhibitor (SERPIN) and MS3_10186 a tetraspanin were selected for *S. haematobium* based on their overall diagnostic performances (**Table 1**). The two proteins were predicted to have a signal peptide attached to them indicating they are utilised outside the cell. MS3_10385 and MS3_10186 were both predicted to be soluble proteins indicating that they are not part of the transmembrane helix. Finally, these two proteins were predicted to be part of the extracellular matrix where immunological reactions are likely to occur.

**Table 1:** Properties and characteristics *S. haematobium* and *S. mansoni* recombinant proteins.

| Description | Protein family | Gene name | Accession number | Source organism | Signal peptide | Transmembrane helix | Cellular localisation |
| --- | --- | --- | --- | --- | --- | --- | --- |
| Neuroserpin | SERPIN | MS3_10385 | A0A095A7Z5 | *S. haematobium* | Signal peptide cleavage site position 27 and 28 | Soluble protein | Extracellular membrane |
| IPSE | Tetraspanin | MS3_10186 | A0A095A2X3 | *S. haematobium* | Signal peptide cleavage site position 20 and 21 | Soluble protein | Extracellular membrane |
| RP26 | SAPLIP | AAB81008 | Q26536 | *S. mansoni* | Signal peptide cleavage site position 18 and 19 | Membrane protein | Extracellular membrane |
| SmSPI | SERPIN | SmSPI | G4LYU6 | *S. mansoni* | Not signal peptide | Soluble protein | Mitochondria |

Saposin-like proteins (SAPLIP) Serine protease inhibitor (SERPIN)

For *S. mansoni* one protein AAB81008 a saposin-like protein (SAPLIP) was selected based on the based overall diagnostic criteria. However, the second protein SmSPI a SERPIN was selected due to its ability to detect early schistosome infections (28) and its potential to detect single worm infections. AAB81008 was predicted to have a signal peptide attached to it and SmSPI was predicted not to possess a signal peptide. AAB81008 was predicted to be a membrane protein and SMSPI was predicted to be a soluble protein. AAB81008 was predicted to be part of the extracellular matrix. SmSPI was predicted to be located in the mitochondria.

### 4.3 Linear B-cell epitopes/peptides

From the 122 peptides *S. haematobium* and *S. mansoni* peptides 40.98 % (n=50) were predicted with ABCpred, 22.95 % (n=28) with Bepi Pred 2, 21.31 % (n=26) and the remaining 14.75 % (n=18) were obtained from literature (**S2 file**). Tables 2 and 3 shows that from the 122 peptides, only 15.57 % were able to distinguish schistosome positives from schistosome negatives or healthy controls.

**Table 2.**
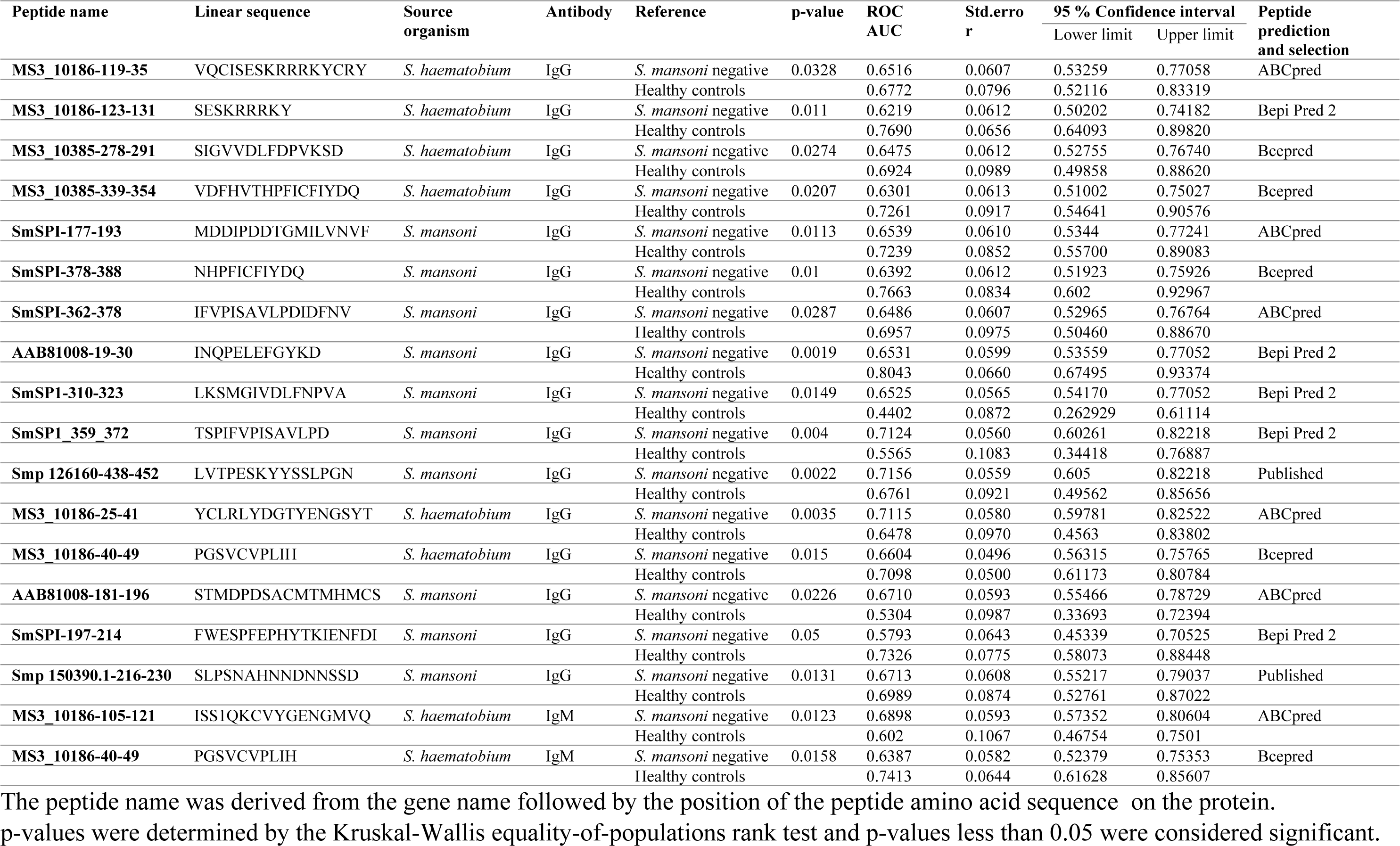
Diagnostic performance of peptides for *S*. *mansoni* IgG and IgM detection.

**Table 3.** Diagnostic performance of peptides for *S. haematobium* IgG and IgM detection.

| Peptide name | Linear sequence | Source organism | Antibody | Reference | p-value | ROC AUC | Std. error | Confidence interval |  | Peptide prediction and selection |
| --- | --- | --- | --- | --- | --- | --- | --- | --- | --- | --- |
|  |  |  |  |  |  |  |  | Lower limit | Upper limit |  |
| **SmSPI-177-193** | MDDIPDDTGMILVNVF | *S. mansoni* | IgG | *S. haematobium* negative | 0.032 | 0.372 | 0.0647 | 0.24602 | 0.49946 | ABCpred |
|  |  |  |  | Healthy controls |  | 0.6302 | 0.1065 | 0.42153 | 0.83894 |  |
| **AAB81008-19-30** | INQPELEFGYKD | *S. mansoni* | IgG | *S. haematobium* negative | 0.0384 | 0.4464 | 0.0666 | 0.31587 | 0.57690 | Bepi Pred 2 |
|  |  |  |  | Healthy controls |  | 0.7326 | 0.0810 | 0.5786 | 0.8916 |  |
| **SmSPI-165-181** | DQQSNGLLEKFFMDDIP | *S. mansoni* | IgG | *S. haematobium* negative | 0.0373 | 0.4092 | 0.0650 | 0.28180 | 0.53668 | Bepi Pred 2 |
|  |  |  |  | Healthy controls |  | 0.6674 | 0.1017 | 0.46811 | 0.86678 |  |
| **AAB81008-19-30** | INQPELEFGYKD | *S. mansoni* | IgM | *S. haematobium* negative | 0.0294 | 0.3692 | 0.0640 | 0.24367 | 0.49470 | Bepi Pred 2 |
|  |  |  |  | Healthy controls |  | 0.6116 | 0.1053 | 0.40522 | 0.81804 |  |

The peptide name was derived from the gene name followed by the position of the peptide amino acid sequence on the protein.
p-values were determined by the Kruskal-Wallis equality-of-populations rank test and p-values less than 0.05 were considered significant.

### 4.4 Diagnostic performance for discriminating *S. mansoni* positives from healthy controls

Out of the 16 peptides that were able to distinguish *S. mansoni* positive sera from schistosome negative or healthy controls sera, only peptide AA81008-19-30 had an excellent diagnostic performance for discriminating *S. mansoni* positives from healthy controls with an AUC value of 0.8043 for IgG peptide microarray (**Fig 1**). Six peptides MS3_10186-123-131, MS3_10385-339-354, SmSPI-177-193, SmSPI-379-388, MS3-10186-40-49 and SmS-197-214 had acceptable diagnostic performances for discriminating *S. mansoni* positives from healthy controls with AUC values ranging from 0.7098 to 0.7763 for IgG peptide microarray (**S3 file**). For IgM peptide microarray the ROC curve analysis for discriminating *S. mansoni* positives from the healthy controls yielded AUC values of 0.602 and 0.7413 for MS3 10186-105-121 and MS3 10186-40-49 respectively (**S3 file**). Peptide MS3 10186-40-49 had acceptable diagnostic performances for discriminating *S. mansoni* positives from healthy controls for both IgG and IgM (**Fig 2**).

**Fig 1:**
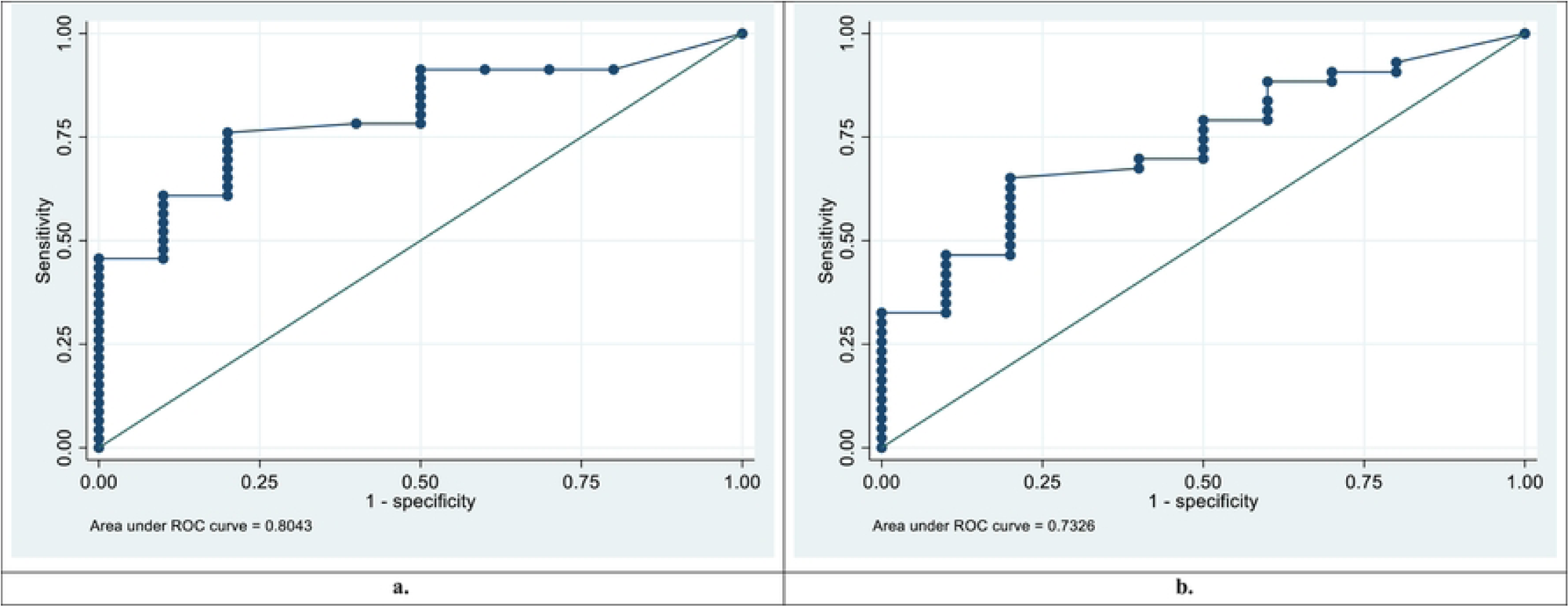
Diagnostic performance for **Peptide AAB81008-19-30 a. Peptide AAB81008-19-30** receiver operating characteristic (ROC) curve and area under the ROC curve (AUC) for discrimination of *S. mansoni* positives from healthy controls for IgG. b. **Peptide AAB81008-19-30** receiver operating characteristic (ROC) curve and area under the ROC curve (AUC) for discrimination of *S. mansoni* positives from healthy controls for IgG.

**Fig 2:**
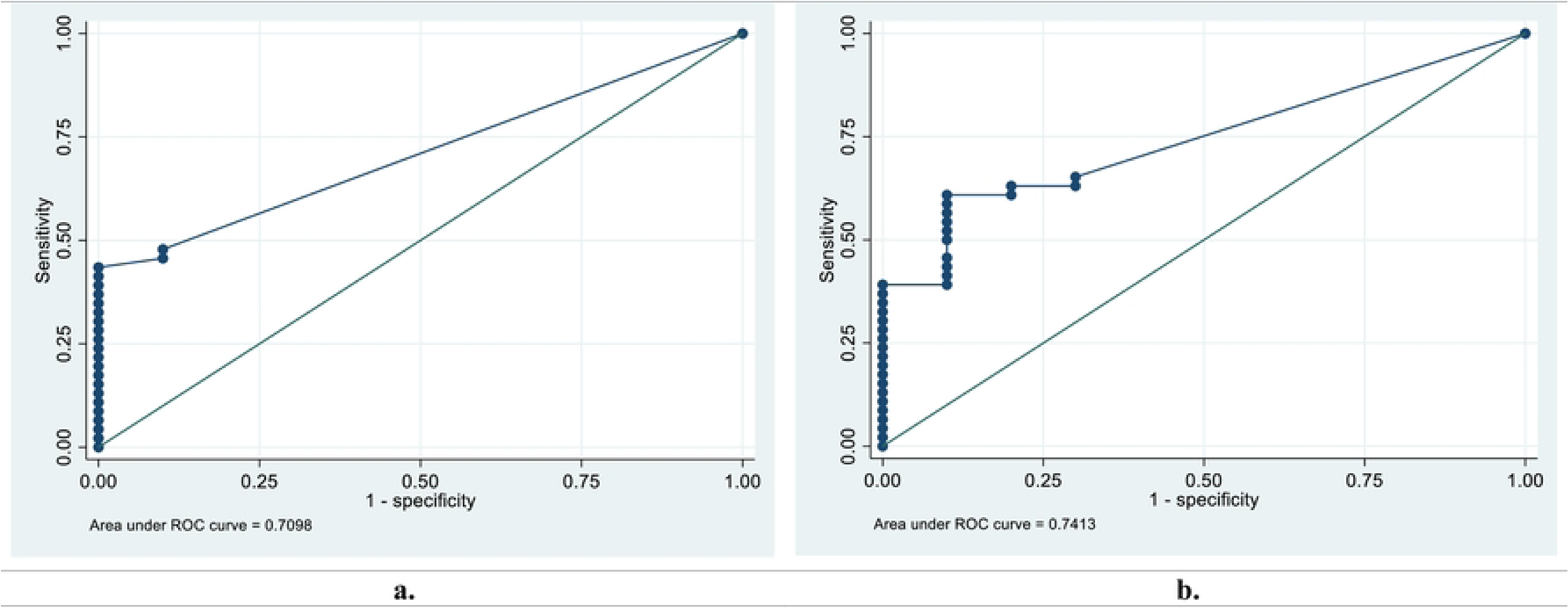
Diagnostic performance for **Peptide MS3 10186-40-49 a. Peptide MS3 10186-40-49** receiver operating characteristic (ROC) curve and area under the ROC curve (AUC) for discrimination of *S. mansoni* positives from healthy controls for IgG. b. **Peptide MS3 10186-40-49** receiver operating characteristic (ROC) curve and area under the ROC curve (AUC) for discrimination of *S. haematobium* positives from healthy controls for IgM.

### 4.5 Diagnostic performance for discriminating *S. mansoni* positives from *S. mansoni* negatives

Three peptides SmSPI-359-372, Smp126160-438-452 and MS3 10186-25-41 had acceptable diagnostic performances for discriminating *S. mansoni* positives from *S. mansoni* negatives with AUC values of 0.7124, 0.7156, 0.7115 respectively for IgG peptide microarray (**fig 3**). For IgM peptide microarray the ROC curve analysis for discriminating the *S. mansoni* positives from the *S. mansoni* negatives yielded AUC values of 0.6898 and 0.6387 for MS3 10186-105-121 and MS3 10186-40-49 respectively (**S3 file**).

**Fig 3:**
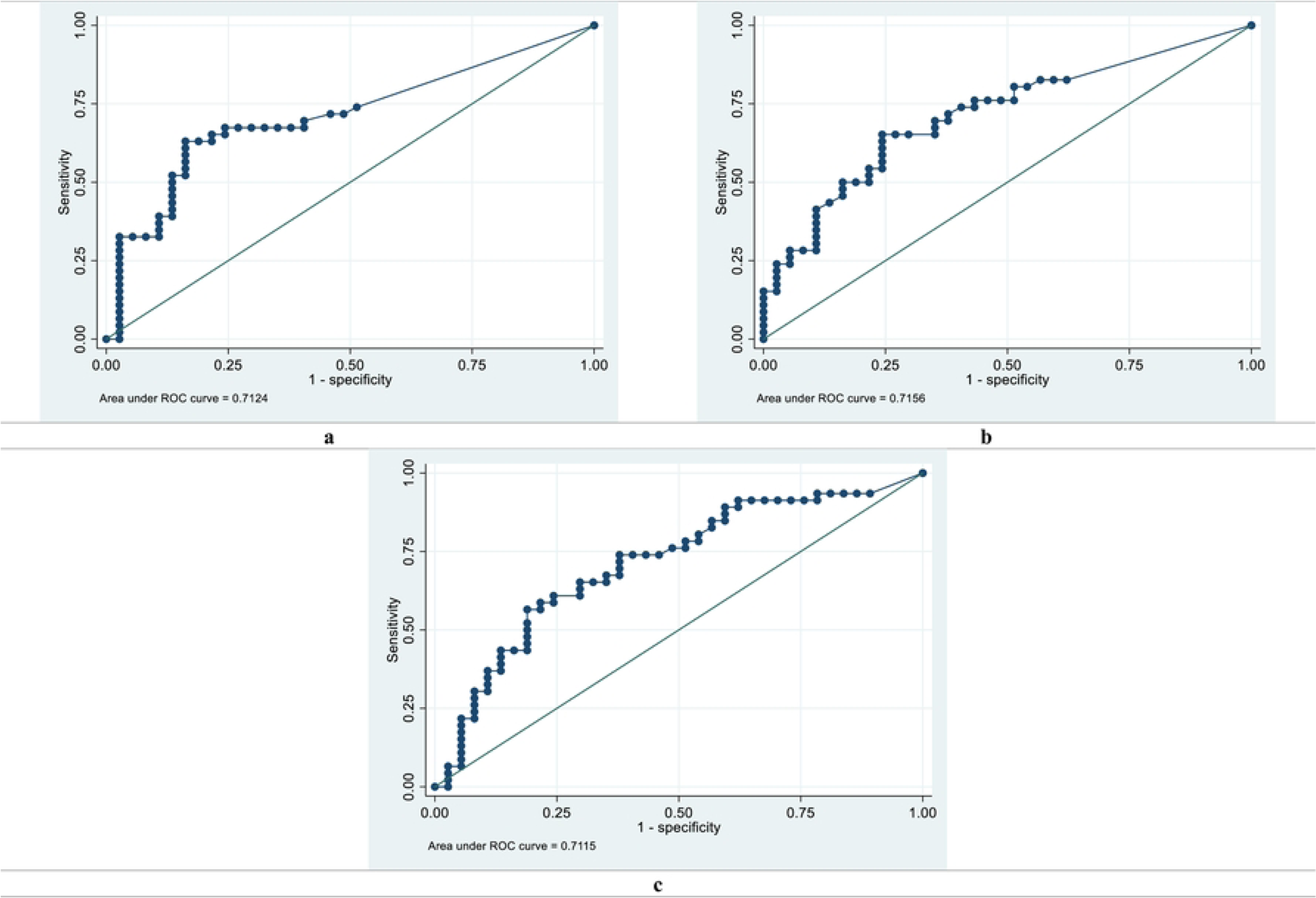
IgG receiver operating characteristic (ROC) curves and area under the ROC curves (AUC) for discriminating *S. mansoni* positives from *S. mansoni* negatives for IgG peptide microarray. a. **Peptide SmpSPI-359-372** b. **Peptide Smp126160-438-452** c. **Peptide MS3 10186-25-41**

### 4.6 Diagnostic performance for discriminating *S. haematobium* positives from *S. haematobium* negatives and healthy controls

The four peptides that were able to distinguish *S. haematobium* positives from *S. haematobium* negatives or healthy controls had inaccurate diagnostic performances for discriminating *S. haematobium* positives from *S. haematobium* negatives for both IgG and IgM peptide microarray (**S4 file**). Peptides SmSPI-177-193 and SmSPI-165-181 had poor diagnostic performances for discriminating *S. haematobium* positives from healthy controls for IgG peptide microarray. Peptide AAB81008 had an acceptable diagnostic performance for discriminating *S. haematobium* positives from healthy controls with an AUC value of 0.7326 for IgG peptide microarray. Peptide AAB81008 was able to discriminate both *S. haematobium* and *S. mansoni* positives from healthy controls (**Fig 1**).

### 4.7 Spatial location of novel peptides with good and acceptable diagnostic performance on the recombinant proteins 3D structures

Understanding the spatial location of peptides within a protein crystal structure is crucial for unravelling the intricacies of molecular interactions and biological functions. In our study DeepView/Swiss-PDB Viewer (www.expasy.org/spdbv/) was used for spatial location of the candidate peptides on the protein crystal structure of recombinant proteins (MS3_10385; MS3_10186; AAB81008 and SmSPI) (**Fig 4**). Peptides AAB1008-19-30, MS3_10186-123-131 and MS3_10385-339-354 were located at the exterior surface of their respective protein structures. Most of the target peptide amino acid sequence was located at the exterior surface of the protein with a few amino acid residues encapsulated within the protein for peptides SmSPI_359-372 and MS3_10186-2541. Lastly, most of the amino acid residues for peptides for SmSPI_177-193, SmSPI_378-388, MS3_10186-40-49 and SmSPI-197-214 were encapsulated within the protein structure and only a few amino acid residues were at the exterior surface of the protein.

**Fig 4:**
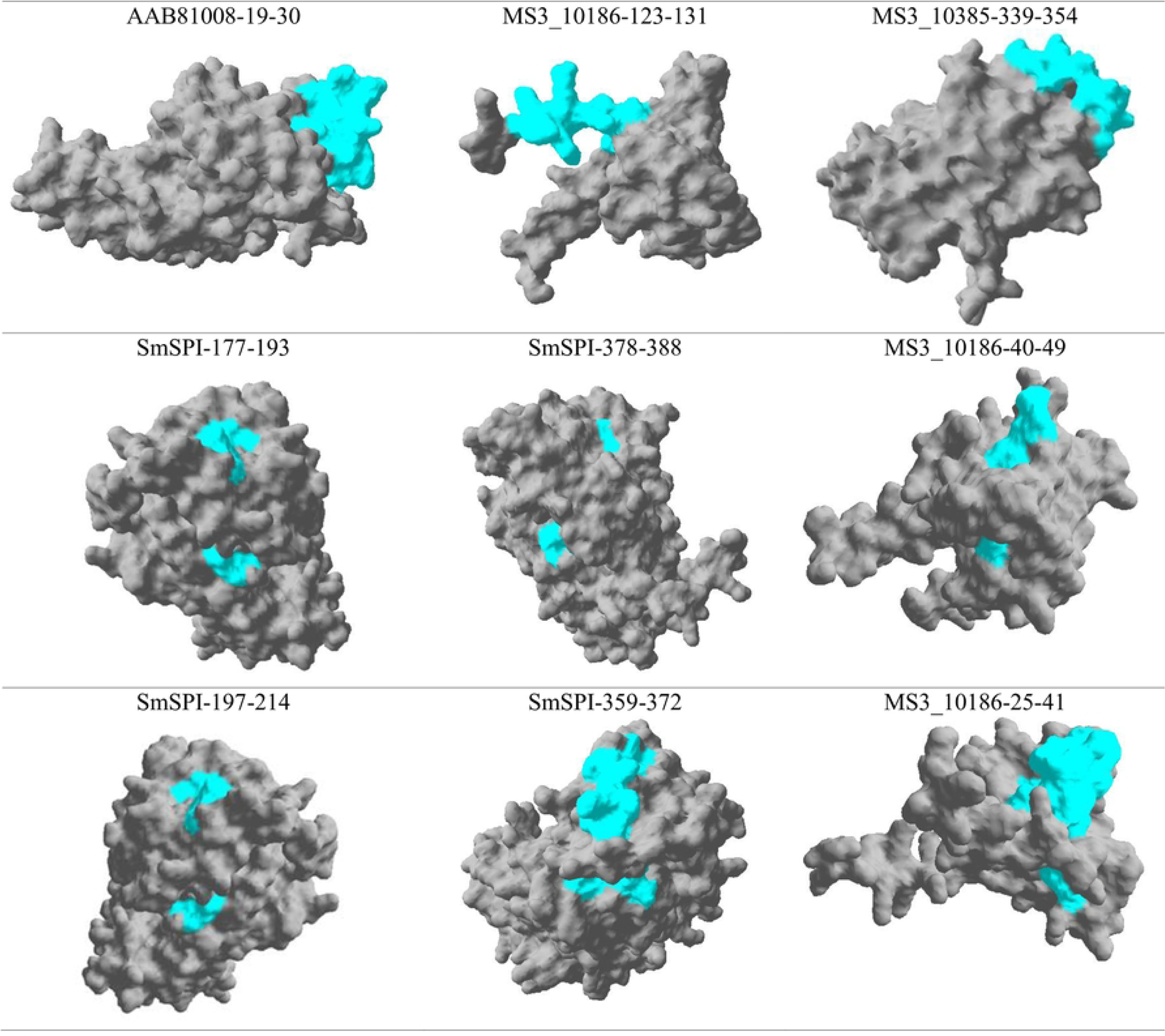
Spatial location of novel peptides with good and acceptable diagnostic performances on the recombinant proteins 3D structures. DeepView/Swiss-PDB Viewer was used to determine the spatial location of the peptides on the crystal structure of recombinant proteins (MS3_10385; MS3_10186; AAB81008 and SmSPI).

## 5 Discussion

As countries target the elimination of schistosomiasis as a public health problem and as the prevalence of the disease decreases due to MDA campaigns, it has become apparently clear that more sensitive field-applicable diagnostics are needed for the effective management and surveillance of schistosome infections as advocated by the WHO (29–31). This is particularly important in low endemicity areas, where microscope-based diagnostic methods may underestimate the true prevalence of the disease (11,32). This background has provided an impetus for this study in the identification of peptides that can be employed in the development of antibody-based diagnostic tools using an immunoinformatic approach and peptide microarray immunoassay validation.

Carvalho and colleagues, 2022 and Lopes and colleagues, 2017 had previously used immunoinformatic approaches and immune-assay validation to identify *S. haematobium* and *S. mansoni* peptides with diagnostic potential (18,19). This study identified one peptide (AA81008-19-30) that had an excellent diagnostic performance for discriminating *S. mansoni* positives from healthy controls for IgG. Six peptides that had acceptable diagnostic performances for discriminating *S. mansoni* positives from healthy controls were identified for IgG. For discriminating *S. mansoni* positives from *S. mansoni* negatives three peptides were identified for IgG. Peptide AAB81008 was shown to have an acceptable diagnostic performance for discriminating *S. haematobium* positives from *S. haematobium* negatives. Peptide AAB81008 was able to discriminate both *S. haematobium* and *S. mansoni* positives from healthy controls.

Eighteen previously published peptides were included on the peptide microarray, 10 from our previous study Vengesai and colleagues, 2022 (26) and 8 peptides from two studies by Carvalho and Colleagues, 2022 and Lopes and Colleagues, 2017 (18,19). Despite Carvalho and Colleagues, 2022 and Lopes and Colleagues, 2017 showing that peptides Sm168240, Smp_136560 (1564–1578), Smp_126160(438–452), Sm140560, Sm041370, Smp_180240(339–353), Smp_150390.1(216–230), Smp_093840(219–233) had AUC values ranging from 0.76 (95 % CI 0.6024-0.9213) to 0.99 (95 % CI 0.987-100) results from this current study show that only one published peptide Smp 126160-438-452 had an acceptable diagnostic performance with an AUC value of 0.7115 (95% CI 0.605-0.82218). The findings may be attributed to the fact that peptide microarray immunoassays were used in the determination of antibody reactivity against the peptides whilst in other two studies (18,19) antibody reactivity was investigated using peptide based serum IgG ELISA. Pertaining the other 10 published peptides XP_035587815.1-269-283, XP_012797374.1-78-92, AAZ29530.1-25-29, XP_012799745.1-16-30, P20287.1-58-72, AAA29903.1-222-237, P09841.3-6-20, AAA29900.1-145-159, P09792.1-29-43, XP_035588858.1-206-220 none of them showed a clear discrimination between the schistosome infected and uninfected groups and these results are in agreement with findings from our previous study (26).

### 5.1 Limitations and recommendations

Ideally, to properly evaluate the performance of the identified peptides their diagnostic accuracy need to be compared to a reference standard test. The reference standard test should be able to discriminate the true positives and true negatives. The absence of a standard reference test or absolute knowledge of the true positives was a significant limitation in the present study (11,33,34). Despite the inappropriateness of doing so, Kato-Katz and urine filtration techniques were used as the reference tests in the current study. The techniques lack sensitivity especially in low endemicity settings; hence it is expected that some positive individuals were misdiagnosed as false-negative (10,11). This might have contributed to the low diagnostic performances of the some identified peptides. To circumvent this limitation, we propose use of the latent class analysis previously described by Mesquita and colleagues, 2022, which combines multiple test results to construct a standard reference outcome (11).

Unfortunately, like most currently available serological tests the peptides identified are not useful for monitoring and evaluation programmes because antibodies to schistosome infections remain detectable long after treatment (35). This limits the clinical value of antibody detection for confirmation of the success of chemotherapy since specific antibodies continue to be present long after the worms have disappeared (14,36). There may be certain antigens (for example peptides) to which certain antibody isotype subclasses like IgG_4_, disappear more rapidly (35). These antibodies can be targeted to circumvent the limitation associated with persisting antibodies. According to the WHO, serological and immunological tests are useful for showing exposure to schistosome infection in people living in non-endemic or low-transmission (37). Alternatively, the identified peptides can be used to develop serological tools for showing exposure to infection for people living in non-endemic and low-transmission areas.

As urogenital and intestinal schistosomiasis are poverty related diseases prevalent in resource-limited settings, cost is a major factor for developing diagnostics for the disease (38). The peptide microarray technology described in this study is too complex and expensive for routine clinical microbiology in resource-limited settings. There it is recommend that the peptides discovered be transferred to a wide range of platforms including, enzyme-linked immunosorbent assay, lateral flow, western blot, and bead-based assays, where they may experdite diagnostics, epidemiology and vaccinology.

### 5.2 Conclusion

In conclusion, 1 peptide with a good diagnostic performance and 9 peptides with acceptable diagnostic performance were identified using the immunoinformatic approach and peptide microarray validation. Identified peptides maybe be used to develop diagnostic tools for showing exposure to schistosome infection in people living in non-endemic or low-transmission areas. Peptides SmPI-177-198 and SmSPI-379-388 may be used to develop a chimeric protein for diagnosis of early *S. mansoni* and single worm infections. However, there is need for validation of the findings with true negative controls from a non-endemic country and a good reference tool.

## Data Availability

All relevant data are within the manuscript and its Supporting Information files.

## 6 Acknowledgments

We thank the communities of Shamva, and Mount Darwin rural districts in the Mashonaland Central province of Zimbabwe for their participation, and support in the study. The authors would like to acknowledge the valuable input of laboratory technicians from the University of Zimbabwe who assisted in sample collection.

## 8 Supplementary files

### S1 file

Excel file

*S. haematobium* and *S. mansoni* demographics and parasitology and peptide microarray immunoassay data sets.

### S2 file

Excel file

*S. haematobium* and *S. mansoni* linear B-cell epitopes

### S3 file

Word document

Receiver operating characteristics (ROC) curve and area under the ROC curve (AUC) for discrimination of *S. mansoni* positives from healthy controls and *S. mansoni* negatives.

### S4 file

Word document

Receiver operating characteristics (ROC) curve and area under the ROC curve (AUC) for discrimination of *S. haematobium* positives from healthy controls and *S. haematobium* negatives.

## References

1. Schistosomiasis [Internet]. [cited 2023 May 24]. Available from: https://www.who.int/news-room/fact-sheets/detail/schistosomiasis

2. Midzi N, Mduluza T, Chimbari MJ, Tshuma C, Charimari L, Mhlanga G, et al. Distribution of Schistosomiasis and Soil Transmitted Helminthiasis in Zimbabwe: Towards a National Plan of Action for Control and Elimination. Kabatereine NB, editor. PLoS Negl Trop Dis [Internet]. 2014 Aug 14 [cited 2020 Mar 3];8(8):e3014. Available from: http://dx.plos.org/10.1371/journal.pntd.0003014

3. Nausch N, Dawson EM, Midzi N, Mduluza T, Mutapi F, Doenhoff MJ. Field evaluation of a new antibody-based diagnostic for Schistosoma haematobium and S. mansoni at the point-of-care in northeast Zimbabwe [Internet]. 2014 [cited 2020 May 20]. Available from: http://www.biomedcentral.com/1471-2334/14/165

4. Vengesai A, Naicker T, Kasambala M, Midzi H, Mduluza-Jokonya T, Rusakaniko S, et al. Clinical utility of peptide microarrays in the serodiagnosis of neglected tropical diseases in sub-Saharan Africa: protocol for a diagnostic test accuracy systematic review. 2021 Jul [cited 2022 Jan 9];11(7):e042279. Available from: https://pubmed.ncbi.nlm.nih.gov/34330850/

5. Archer J, Barksby R, Pennance T, Rostron P, Bakar F, Knopp S, et al. Analytical and Clinical Assessment of a Portable, Isothermal Recombinase Polymerase Amplification (RPA) Assay for the Molecular Diagnosis of Urogenital Schistosomiasis. Mol 2020, Vol 25, Page 4175 [Internet]. 2020 Sep 11 [cited 2023 May 24];25(18):4175. Available from: https://www.mdpi.com/1420-3049/25/18/4175/htm

6. Lim MD, Brooker SJ, Belizario VY, Gay-Andrieu F, Gilleard J, Levecke B, et al. Diagnostic tools for soil-transmitted helminths control and elimination programs: A pathway for diagnostic product development. PLoS Negl Trop Dis [Internet]. 2018 Mar 1 [cited 2023 May 24];12(3):e0006213. Available from: https://journals.plos.org/plosntds/article?id=10.1371/journal.pntd.0006213

7. Mu Y, Gordon CA, Olveda RM, Ross AG, Olveda DU, Marsh JM, et al. Identification of a linear B-cell epitope on the Schistosoma japonicum saposin protein, SjSAP4: Potential as a component of a multi-epitope diagnostic assay. PLoS Negl Trop Dis [Internet]. 2022 [cited 2023 May 24];16(7):e0010619. Available from: https://journals.plos.org/plosntds/article?id=10.1371/journal.pntd.0010619

8. Hoermann J, Kuenzli E, Schaefer C, Paris DH, Bühler S, Odermatt P, et al. Performance of a rapid immuno-chromatographic test (Schistosoma ICT IgG-IgM) for detecting Schistosoma-specific antibodies in sera of endemic and non-endemic populations. PLoS Negl Trop Dis [Internet]. 2022 [cited 2023 May 24];16(5):e0010463. Available from: https://journals.plos.org/plosntds/article?id=10.1371/journal.pntd.0010463

9. Menezes DL, Santos CT de J, Oliveira YLDC, Campos VTC, Negrão-Corrêa DA, Geiger SM, et al. Accuracy Study of Kato-Katz and Helmintex Methods for Diagnosis of Schistosomiasis Mansoni in a Moderate Endemicity Area in Sergipe, Northeastern Brazil. Diagnostics 2023, Vol 13, Page 527 [Internet]. 2023 Jan 31 [cited 2023 May 24];13(3):527. Available from: https://www.mdpi.com/2075-4418/13/3/527/htm

10. Colley DG, King CH, Kittur N, Ramzy RMR, Secor WE, Fredericks-James M, et al. Evaluation, Validation, and Recognition of the Point-of-Care Circulating Cathodic Antigen, Urine-Based Assay for Mapping Schistosoma mansoni Infections. Am J Trop Med Hyg [Internet]. 2020 May 12 [cited 2023 May 24];103(1_Suppl):42–9. Available from: https://www.ajtmh.org/view/journals/tpmd/103/1_Suppl/article-p42.xml

11. Mesquita SG, Caldeira RL, Favre TC, Massara CL, Beck LCNH, Simões TC, et al. Assessment of the accuracy of 11 different diagnostic tests for the detection of Schistosomiasis mansoni in individuals from a Brazilian area of low endemicity using latent class analysis. Front Microbiol. 2022 Dec 15;13:4683.

12. Pearson MS, Tedla BA, Mekonnen GG, Proietti C, Becker L, Nakajima R, et al. Immunomics-guided discovery of serum and urine antibodies for diagnosing urogenital schistosomiasis: a biomarker identification study. The Lancet Microbe [Internet]. 2021 Nov 1 [cited 2023 May 26];2(11):e617–26. Available from: http://www.thelancet.com/article/S2666524721001506/fulltext

13. Imai N, Rujeni N, Nausch N, Bourke CD, Appleby LJ, Cowan G, et al. Exposure, infection, systemic cytokine levels and antibody responses in young children concurrently exposed to schistosomiasis and malaria. Parasitology. 2011 Oct;138(12):1519–33.

14. Ogongo P, Kariuki TM, Wilson RA. Diagnosis of schistosomiasis mansoni: an evaluation of existing methods and research towards single worm pair detection. Parasitology [Internet]. 2018 Sep 1 [cited 2022 Mar 12];145(11):1355–66. Available from: https://www.cambridge.org/core/journals/parasitology/article/abs/diagnosis-of-schistosomiasis-mansoni-an-evaluation-of-existing-methods-and-research-towards-single-worm-pair-detection/552001BF61F171FB60F11577DC42280A

15. Ma L, Zhao W, Hou X, Liu M, Li Y, Shen L, et al. Identification of linear epitopes in SjSP-13 of Schistosoma japonicum using a GST-peptide fusion protein microplate array. Parasites and Vectors [Internet]. 2019 Oct 30 [cited 2020 Nov 22];12(1). Available from: https://pubmed-ncbi-nlm-nih-gov.ukzn.idm.oclc.org/31666115/

16. Falconi-Agapito F, Kerkhof K, Merino X, Bakokimi D, Torres F, Van Esbroeck M, et al. Peptide Biomarkers for the Diagnosis of Dengue Infection. Front Immunol. 2022 Jan 26;13:52.

17. de Oliveira EJ, Kanamura HY, Takei K, Hirata RDC, Valli LCP, Nguyen NY, et al. Synthetic peptides as an antigenic base in an ELISA for laboratory diagnosis of schistosomiasis mansoni. Trans R Soc Trop Med Hyg [Internet]. 2008 Apr 1 [cited 2022 Feb 15];102(4):360–6. Available from: https://pubmed.ncbi.nlm.nih.gov/18314149/

18. Carvalho GBF, Resende DM, Siqueira LMV, Lopes MD, Lopes DO, Coelho PMZ, et al. Selecting targets for the diagnosis of Schistosoma mansoni infection: An integrative approach using multi-omic and immunoinformatics data. PLoS One [Internet]. 2017 Aug 1 [cited 2022 Jan 9];12(8):e0182299. Available from: https://journals.plos.org/plosone/article?id=10.1371/journal.pone.0182299

19. Lopes MD, Oliveira FM, Coelho IEV, Passos MJF, Alves CC, Taranto AG, et al. Epitopes rationally selected through computational analyses induce T-cell proliferation in mice and are recognized by serum from individuals infected with Schistosoma mansoni. Biotechnol Prog [Internet]. 2017 May 1 [cited 2022 Feb 14];33(3):804–14. Available from: https://pubmed.ncbi.nlm.nih.gov/28371522/

20. Van Regenmortel MHV. Structural and functional approaches to the study of protein antigenicity [Internet]. Vol. 10, Immunology Today. Immunol Today; 1989 [cited 2021 Jun 14]. p. 266–72. Available from: https://pubmed.ncbi.nlm.nih.gov/2478146/

21. Giacò L, Amicosante M, Fraziano M, Gherardini PF, Ausiello G, Helmer-Citterich M, et al. B-Pred, a structure based B-cell epitopes prediction server. Adv Appl Bioinforma Chem [Internet]. 2012 Jul 25 [cited 2021 Jun 14];5(1):11–21. Available from: 10.2147/AABC.S30620

22. Vengesai A, Kasambala M, Mutandadzi H, Mduluza-Jokonyaid TL, Mduluzaid T, Naicker T, et al. Scoping review of the applications of peptide microarrays on the fight against human infections. PLoS One [Internet]. 2022 Jan [cited 2022 Feb 13];17(1):e0248666. Available from: https://journals.plos.org/plosone/article?id=10.1371/journal.pone.0248666

23. Sanchez-Lockhart M, Reyes DS, Gonzalez JC, Garcia KY, Villa EC, Pfeffer BP, et al. Qualitative Profiling of the Humoral Immune Response Elicited by rVSV-ΔG-EBOV-GP Using a Systems Serology Assay, Domain Programmable Arrays. Cell Rep [Internet]. 2018 Jul 24 [cited 2020 Nov 26];24(4):1050–1059.e5. Available from: 10.1016/j.celrep.2018.06.077

24. Sanchez-Trincado JL, Gomez-Perosanz M, Reche PA. Fundamentals and Methods for T- and B-Cell Epitope Prediction [Internet]. Vol. 2017, Journal of Immunology Research. Hindawi Limited; 2017 [cited 2021 Jan 14]. Available from: https://pubmed.ncbi.nlm.nih.gov/29445754/

25. Vengesai A, Muleya V, Midzi H, Tinago TV, Chipako I, Manuwa M, et al. Diagnostic performances of Schistosoma haematobium and Schistosoma mansoni recombinant proteins, peptides and chimeric proteins antibody based tests. Systematic scoping review. PLoS One. 2023 Mar 1;18(3 March).

26. Vengesai A, Naicker T, Midzi H, Kasambala M, Mduluza-Jokonya TL, Rusakaniko S, et al. Multiplex peptide microarray profiling of antibody reactivity against neglected tropical diseases derived B-cell epitopes for serodiagnosis in Zimbabwe. PLoS One [Internet]. 2022 Jul 1 [cited 2023 Jan 31];17(7):e0271916. Available from: https://journals.plos.org/plosone/article?id=10.1371/journal.pone.0271916

27. Vengesai A, Naicker T, Midzi H, Kasambala M, Muleya V, Chipako I, et al. Peptide microarray analysis of in-silico predicted B-cell epitopes in SARS-CoV-2 sero-positive healthcare workers in Bulawayo, Zimbabwe. Acta Trop [Internet]. 2023 Feb 1 [cited 2023 Jan 31];238:106781. Available from: https://linkinghub.elsevier.com/retrieve/pii/S0001706X22004727

28. Tanigawa C, Fujii Y, Miura M, Nzou SM, Mwangi AW, Nagi S, et al. Species-Specific Serological Detection for Schistosomiasis by Serine Protease Inhibitor (SERPIN) in Multiplex Assay. PLoS Negl Trop Dis [Internet]. 2015 [cited 2020 May 20];9(8):e0004021. Available from: https://journals.plos.org/plosntds/article?id=10.1371/journal.pntd.0004021

29. Colley DG, King CH, Kittur N, Ramzy RMR, Secor WE, Fredericks-James M, et al. Evaluation, Validation, and Recognition of the Point-of-Care Circulating Cathodic Antigen, Urine-Based Assay for Mapping Schistosoma mansoni Infections. Am J Trop Med Hyg [Internet]. 2020 May 12 [cited 2023 Dec 4];103(1_Suppl):42–9. Available from: https://www.ajtmh.org/view/journals/tpmd/103/1_Suppl/article-p42.xml

30. Archer J, Barksby R, Pennance T, Rostron P, Bakar F, Knopp S, et al. Analytical and Clinical Assessment of a Portable, Isothermal Recombinase Polymerase Amplification (RPA) Assay for the Molecular Diagnosis of Urogenital Schistosomiasis. Mol 2020, Vol 25, Page 4175 [Internet]. 2020 Sep 11 [cited 2023 Dec 4];25(18):4175. Available from: https://www.mdpi.com/1420-3049/25/18/4175/htm

31. Pearson MS, Tedla BA, Mekonnen GG, Proietti C, Becker L, Nakajima R, et al. Immunomics-guided discovery of serum and urine antibodies for diagnosing urogenital schistosomiasis: a biomarker identification study. The Lancet Microbe [Internet]. 2021 Nov 1 [cited 2022 Feb 14];2(11):e617–26. Available from: http://www.thelancet.com/article/S2666524721001506/fulltext

32. Nausch N, Dawson EM, Midzi N, Mduluza T, Mutapi F, Doenhoff MJ. Field evaluation of a new antibody-based diagnostic for Schistosoma haematobium and S. mansoni at the point-of-care in northeast Zimbabwe. BMC Infect Dis [Internet]. 2014 Mar 26 [cited 2023 May 27];14(1):1–9. Available from: https://bmcinfectdis.biomedcentral.com/articles/10.1186/1471-2334-14-165

33. Torlakovic EE, Francis G, Garratt J, Gilks B, Hyjek E, Ibrahim M, et al. Standardization of negative controls in diagnostic immunohistochemistry: Recommendations from the international Ad Hoc expert panel. Appl Immunohistochem Mol Morphol [Internet]. 2014 [cited 2022 May 10];22(4):241–52. Available from: https://journals.lww.com/appliedimmunohist/Fulltext/2014/04000/Standardization_of_Negative_Controls_in_Diagnostic.1.aspx

34. West R, Kobokovich A. Understanding the Accuracy of Diagnostic and Serology Tests: Sensitivity and Specificity Factsheet. 2020;

35. Diagnostic target product profiles for monitoring, evaluation and surveillance of schistosomiasis control programmes [Internet]. [cited 2022 Jan 28]. Available from: https://www.who.int/publications/i/item/9789240031104

36. Ogongo P. Identification and evaluation of Schistosoma mansoni proteins as diagnostic targets for schistosomiasis. Int J Infect Dis. 2014 Apr 1;21:365.

37. Schistosomiasis [Internet]. [cited 2023 Dec 4]. Available from: https://www.who.int/news-room/fact-sheets/detail/schistosomiasis

38. Rivera J, Mu Y, Gordon CA, Jones MK, Cheng G, Cai P. Current and upcoming point-of-care diagnostics for schistosomiasis. Trends Parasitol [Internet]. 2023 Nov 23 [cited 2023 Dec 4]; Available from: https://linkinghub.elsevier.com/retrieve/pii/S1471492223002805

